# In Vivo Intralesional Positronium Lifetime Imaging of Thyroid Cancer Using Clinically Routine I-124 PET/CT

**DOI:** 10.1101/2025.05.28.25328504

**Authors:** Lorenzo Mercolli, William M. Steinberger, Tilman Läppchen, Michelle Amon, Carola Bregenzer, Maurizio Conti, Ângelo R. Felgosa Cardoso, Clemens Mingels, Paweł Moskal, Narendra Rathod, Hasan Sari, Ewa Ł. Stępień, Sabine Weidner, Axel Rominger, Kuangyu Shi, Robert Seifert

## Abstract

**Purpose:** Measuring the lifetime of orthopositronium (oPs), the spin triplet of an electron and positron, has emerged as a promising approach for assessing tumor microenvironment characteristics, leveraging the susceptibility of oPs lifetime to local molecular factors such as the oxygenation level. This study investigates the feasibility of intralesional oPs lifetime measurements in patients with thyroid cancer using a commercial long axial field-of-view (LAFOV) PET/CT scanner.

**Methods:** Three patients with thyroid cancer underwent two PET/CT scans on a Biograph Vision Quadra (Siemens Healthineers, Knoxville TN, USA) at 2 and 26 hours post-administration [^124^I]NaI, which included a 30 min scan in singles mode (a prototype feature for saving single crystal interactions without coincidence sorting) that allows to identify three-photon events. For regions with high ^124^I uptake, we determined the lifetime of oPs together with Bayesian errors estimation for a voxel-based (with a voxel size of 7 × 7 × 7 mm^3^), lesion based and organ based analysis.

**Results:** The oPs lifetime of the voxel with the highest count statistics and activity concentration of 108.1 kBq mL^−1^ is 1.77 ± 0.26 ns. The visualization of the oPs lifetimes inside the tumor lesion was feasible, though it remains affected by statistical error. On a per-lesion basis, the lifetime could be measured (exemplary lesion:1.90 ± 0.17 ns) with mean activity concentrations ranging from 9.5 to 34.1 kBq mL^−1^ and volumes 1.31 to 6.27 kBq mL^−1^. The positronium lifetime measurement error shows a slight correlation (*r* =0.48 with *BF* _10_=7.88) with activity concentration of the regarded region. Besides lesions, the positronium lifetime could also be determined in several organs (average oPs lifetime in exemplary organs: liver 1.68 ± 0.08 ns, left atrium 1.83 ± 0.17 ns, right parotid gland: 2.24 ± 0.12 ns, left kidney 1.87 ± 0.12 ns).

**Conclusion:** This is the first report human in-vivo measurement of oPs lifetime with [^124^I]NaI using a commercial LAFOV PET/CT scanner. The voxel-wise oPs lifetime imaging is feasible, though with a high statistical uncertainty. Lesion-based measurements can yield satisfactory statistical precision even for small volumes under clinical conditions. Future studies can leverage our approach to analyze the tumor microenvironment with regard to oPs lifetime as a surrogate marker for oxygenation.

## 1 Introduction

Conventional positron emission tomography (PET) traditionally relies on the detection of two 511 keV annihilation photons to map the spatial distribution of radiotracers in the human body. Recently, after invention of positronium imaging [1, 2],there has been significant interest to go beyond standard coincidence PET imaging by measuring the lifetime of orthopositronium (oPs) - a bound state of an electron and a positron with spin 1 - in human tissue [1, 3–18]. The lifetime of oPs shortens to a few nanoseconds due to interaction with surrounding tissue. This dependence on the local molecular environment makes oPs lifetime measurements a tool for assessing microenvironmental features and potentially provide an additional diagnostic information.

Last year, Ref. [19] presented the first patient study using the prototype scanner J-PET. Refs. [20–23] showed that in vivo oPs lifetime measurements are possible on a commercial long axial field-of-view (LAFOV) PET/CT. However, measuring oPs lifetimes in a clinical setting has proven challenging. Measuring the oPs lifetime through positron annihilation lifetime spectroscopy (PALS) (see e.g. Refs. [11, 12, 24] and references therein) requires the detection of two annihilation photons and a prompt photon from the same nuclear decay/deexcitation, i.e. three-photon events (3*γ*E). Furthermore, non-pure positron emitting radionuclides that are used in clinical routine have rather small prompt-photon branching ratios (BR) since in standard PET imaging, the prompt photons are considered nuisance that needs to be corrected (see e.g. Ref. [25]). Despite the increased sensitivity that long axial field-of-view PET/CT scanners provide [26, 27], the methodology of Ref. [20] for selecting 3*γ*E in conjunction with the restricted energy range of the scanner available to Refs. [20–23] is limited by count statistics. From these studies, ^124^I has crystallized as the most promising candidate for PALS measurements with a commercial PET/CT due to its prompt photon energy of 602.73 ± 0.08 keV with a reasonably high BR of 62.9 ± 0.7 % (which reduces to 12.0 ± 1.1 % when electron capture decays are removed).

In Refs. [1, 4] positronium lifetime imaging (oPsLI), i.e. determining the oPs life-time for single voxels in an image, was proposed. The image reconstruction process for oPsLI has triggered substantial research and alternative event selection and more refined image reconstruction techniques (see e.g. Refs. [10, 16–18, 28–30] may in the future alleviate the limitation of the current methodology form Refs. [20, 21].

In a phantom study in Ref. [22], we showed that oPsLI is feasible ^124^I on a commercial PET/CT scanner using the methodology of Ref. [20]. As shown in Refs. [20–23], the Biograph Vision Quadra (Siemens Healthineers, Knoxville TN, USA) can optimally exploit ^124^I’s physical properties, which is not the case for high-energy prompt photon emitting radionuclides like ^68^Ga, ^82^Rb or ^44^Sc. Apart from the technical capabilities of Quadra, the differentiation of thyroid cancer may be associated with tumor hypoxia [31, 32] and therefore has the potential to provide an excellent clinical application for oPsLI. In this study, we therefore investigate oPsLI thyroid cancer patients using ^124^I. Through an early and a late scan of three patients, we investigate the oPs lifetime in organs and tumor lesions. Despite low statistical precision, we construct oPsLI for the most emblematic cases.

## 2 Materials and methods

This study included three patients with thyroid carcinoma who have been referred to ^131^I-therapy for initial ablation after thyroidectomy (S1) or because of recurrent thyroid cancer (S2 and S3) with rising thyreoglobulin levels. The [124I] PETs were performed as part of routine clinical practice for staging and to assess the extent of postsurgical remnant tissue (S1) and/or to plan the [131I] therapy dose (S2, S3). Patients presented with follicular thyroid cancer and cervical lymph node metastases (S1), poorly differentiated thyroid cancer with a bone metastasis that showed intense uptake on [^18^F]FDG (S2), and papillary thyroid cancer with a recently resected lung metastasis (S3). Data analysis was done retrospectively.

The three patients received an oral capsule of ^124^I sodium iodide (3D Imaging LLC, Little Rock, AR, USA). The administered activity was 41.02 MBq, 40.11 MBq and 39.44 MBq, respectively, and the patients were scanned twice at 2 h and at 26 h post application on a Biograph Vision Quadra (Siemens Healthineers, Knoxville TN, USA) LAFOV PET/CT. The early and late scan followed the same protocol, i.e. a PET scan in coincidence mode for 15 min combined with a 30 min scan for the PALS measurement. The two-photon, i.e. standard coincidence, images were reconstructed with the vendor’s TrueX reconstruction algorithm in ultra high sensitivity mode, including point-spread-function, attenuation, relative scatter and prompt photon correction and with a 440 × 440 matrix, 4 iterations, 5 subsets. For the PALS measurement, the scanner was operated in singles acquisition mode, a prototype feature to record every single-crystal interaction without the conventional two-photon coincidence sorting.

From the singles list-mode data, 3*γ*E were identified using an offline prototype software. The detailed methodology is described in Ref. [20]. For the 3*γ*E selection, the energy window for the annihilation photons is 460 keV to 545 keV and for the prompt photon 568 keV to 639 keV. The localization of the 3*γ*E relies solely on the time-of-flight (TOF) information from the annihilation photons, i.e. no reconstruction algorithm like in Refs. [10, 16–18, 28, 33]) used. The time window for the two annihilation photons is 4.2 ns. Spatial cuts, i.e. a minimal crystal distance, were also used to suppress random events arising from ^176^Lu in Quadra’s crystals.

Segmentations of volumes-of-interest (VOI) were performed either on the CT or coincidence PET images using LIFEx and TotalSegmentator [34] and checked and/or amended by a nuclear medicine physician. The six lesions of S1 were are clearly visible in the histoimage (number of detected 3*γ*E as voxel value). The other patients did not show regions with pathological uptake in the late scans. For the organ-level oPs lifetime determination, we considered the heart chambers, parotid glands, kidneys and liver in the early scans of all three patients. In the late scans, only the parotid glands and five lesions of S1 show enough 3*γ*E to be considered for *τ*_3_ fitting.

oPsLIwas constructed with a voxel size of 7 × 7 × 7 mm^3^ on the late scan of S1 with a focus on the lesions and the early scan for S2. Though highly desirable from a clinical perspective, smaller voxel sizes do not seem to be very meaningful given the large positron range of ^124^I (about 10 mm in water [35]) and the absence of a true reconstruction algorithm for 3*γ*E localization in our methodology. PALS were generated by histogramming the measured delay between the prompt photon and the annihilation photons for each 3*γ*E in a VOI or single voxel, as described in Ref. [20].

A three-component model was applied for fitting the measured PALS, consisting of exponential decay terms for direct annihilation and pPs, and oPs contribution

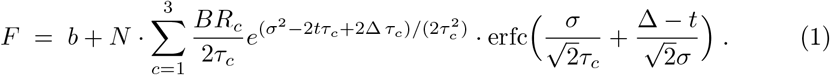

In Eq. (1), *b* denotes the background and *N* is a normalization constant. *BR*_*c*_ with *c* = 1 … 3 and ∑ _*c*_ *BR*_*c*_ = 1 are the relative branching ratios of pPs, direct annihilation and oPs, respectively. *σ* and Δ are the parameters of the Gaussian, which models the whole measurement chain and is convoluted with the three lifetime components. For an accurate estimate of the statistical uncertainties, our analysis relies on a Bayesian framework for the fitting of PALS single VOI or voxels outlined in Refs. [20–22]. The priors on the fitting parameters are the same as in Ref. [22], i.e.

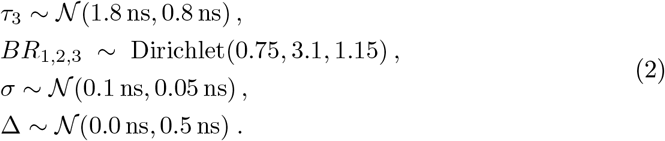

The background *b* is fixed to the mean of the 3*γ*E with time differences *<* 2 ns, while the fit is performed on time differences between ™ 1.5 to 8.0 ns. We checked that there is no significant impact on the posterior distribution of *τ*_3_ whether *τ*_1,2_ are fixed to literature values or considered as fit parameters with strong priors [22]. All errors are reported in terms of marginalized posterior distributions. While for *τ*_3_ it is very similar to a Gaussian distribution, and hence quoting a standard deviation is acceptable, the relative component’s BR need to be reported in terms of 68% highest-density-intervals (HDI).

## 3 Results

Fig. 1 shows the oPsLI of two coronal and axial slices in the region of the lesions. *τ*_3_ and the statistical error Δ*τ*_3_ were determined for 7 × 7 × 7 mm^3^ voxels. The voxel size of the coincidence PET image is 1.9 × 1.9 × 1.65 mm^3^. In order to improve the visualization, in Fig. 1 the oPsLI was resampled and interpolated to the resolution of the coincidence PET image. Only voxels with a relative standard deviation *<* 65% on 3*γ*E with time differences *<* − 2 ns were considered for *τ*_3_ fitting since only few voxels show a relative standard deviation of less than 20% which would allow for a small Δ*τ*_3_. The PALS of the two voxels with the highest number of 3*γ*E are shown in the bottom row of Fig. 1.

**Fig. 1:**
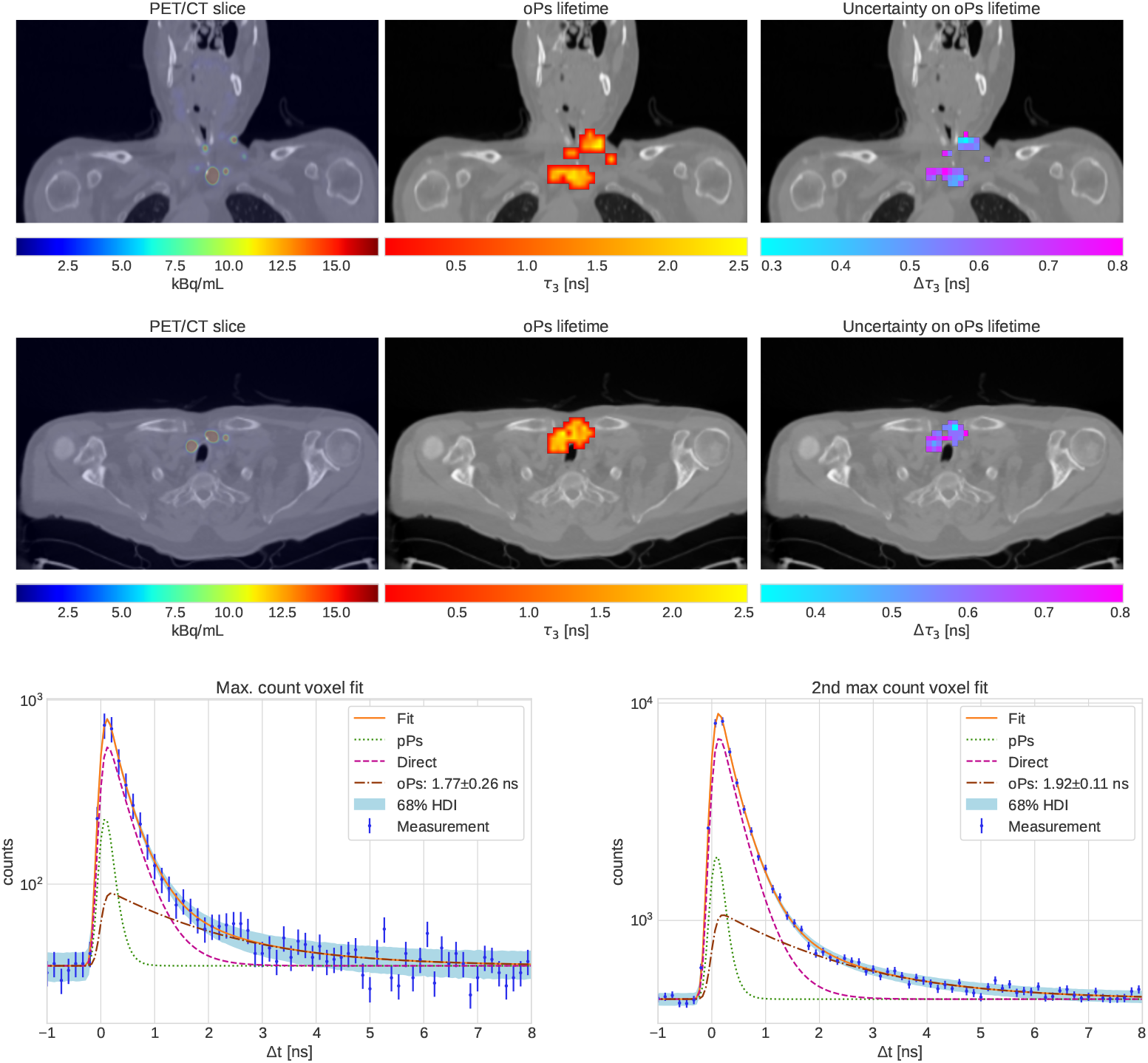
oPsLI of coronal (*top row*) and axial (*center row*) slices of S1 in the region with lesions together with the coincidence PET slices (*left*) and the error voxel-wise error on *τ*_3_ (*right*). For improved visualization the *τ*_3_ image was resampled and interpolated on the coincidence PET image resolution. The *bottom row* shows the PALS of the two voxels with the highest count statistics.

The early scans of the three patients showed, unsurprisingly, most uptake in the gastrointestinal tract. With respect to oPsLI this is not particularly interesting. Nevertheless, Fig. 2 shows the oPsLI of S1 at 2 h post application. Only voxels with a relative standard deviation *<* 60% on 3*γ*E with time differences *<* − 2 ns were considered for *τ*_3_ fitting.

**Fig. 2:**
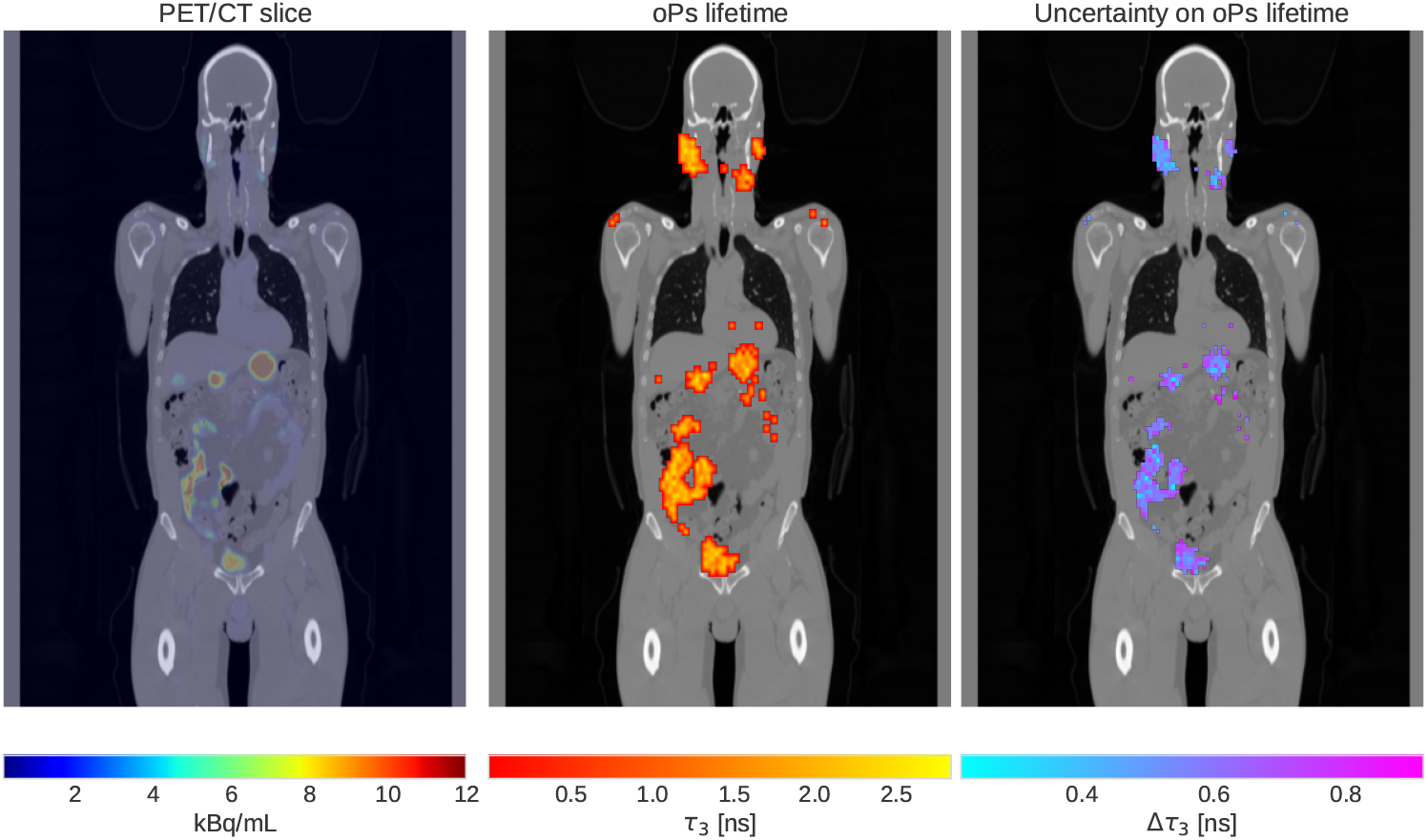
A oPsLI coronal slice of S2’s 2 h scan (*center*) together with the corresponding error on *τ*_3_ (*right*) and coincidence PET image (*left*).

Focusing our attention to the VOI-based analysis, Tab. 1 summarizes the lesions’ properties, i.e. volume, mean voxel value within the VOI of the coincidence PET image and SUV_max_, and if the lesion can be delineated on the FDG scan. The last column shows the oPs lifetime of the lesions. Clearly, the statistical uncertainty on *τ*_3_ correlates with the mean activity concentration in the lesion (see also bottom right panel of Fig. 4). In Fig. 3 we visualize the PALS of the lesions L1 to L4. L5 and L6 have the lowest count statistics and the PALS would be even worse than for L1. It should be noted that that the PALS are shown on a log scale and that the volume of the lesions under consideration is rather small. Finally, the top panel Fig. 4 shows the PALS of all 3*γ*E that were identified within the five lesions. As a reference, Tab. 2 ns reports the fit results for several VOI in the early scans of all three patients.

**Fig. 3:**
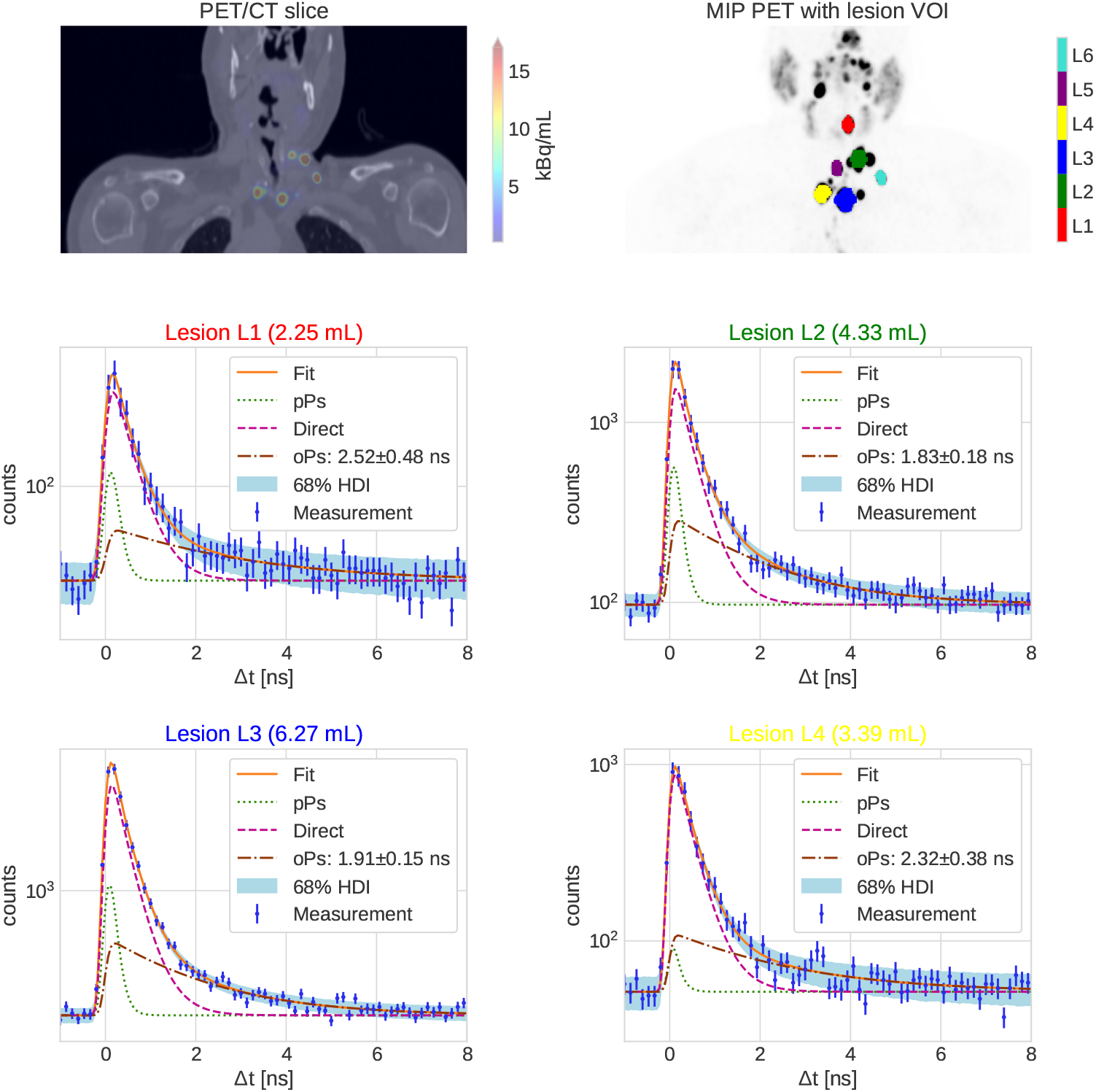
The *top row* shows a slice of the coincidence PET/CT (*left*) and the MIP with the segmentation of the five lesions (*right*). The *center* and *bottom row* show PALS of L1 to L4 of S1 at 26 h post application.

**Fig. 4:**
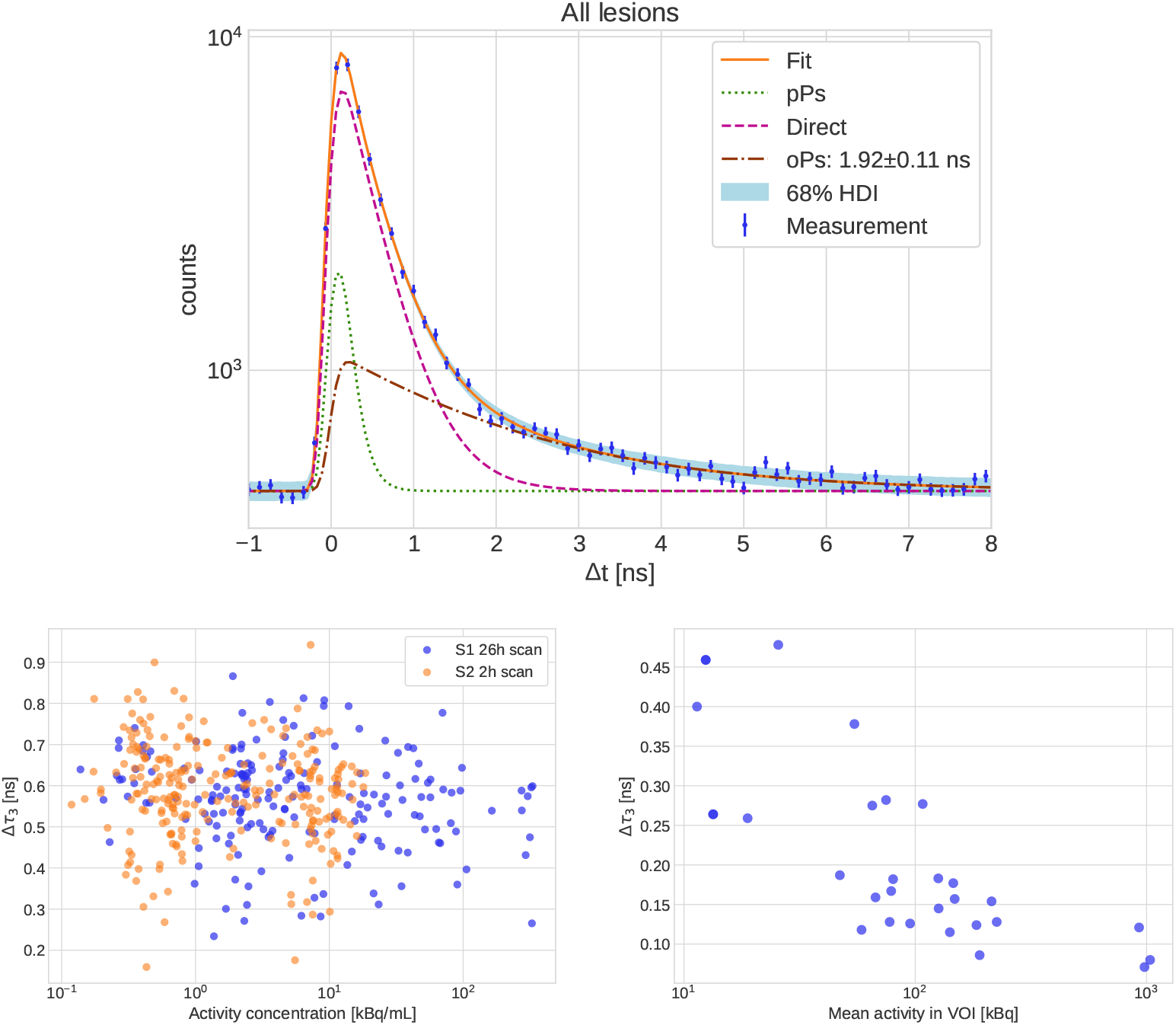
*Top panel:* PALS of all 3*γ*E within the lesions L1 to L5. Statistical uncertainty Δ*τ*_3_ as a function of the max. activity concentration in 7× 7 ×7 mm^3^ voxels for S1’s 26 h scan and S2’s 2 h scan (*bottom left*) or as a function of mean activity in a single VOI (*bottom right*) for the VOI listed in Tab. 2.

**Table 1:**
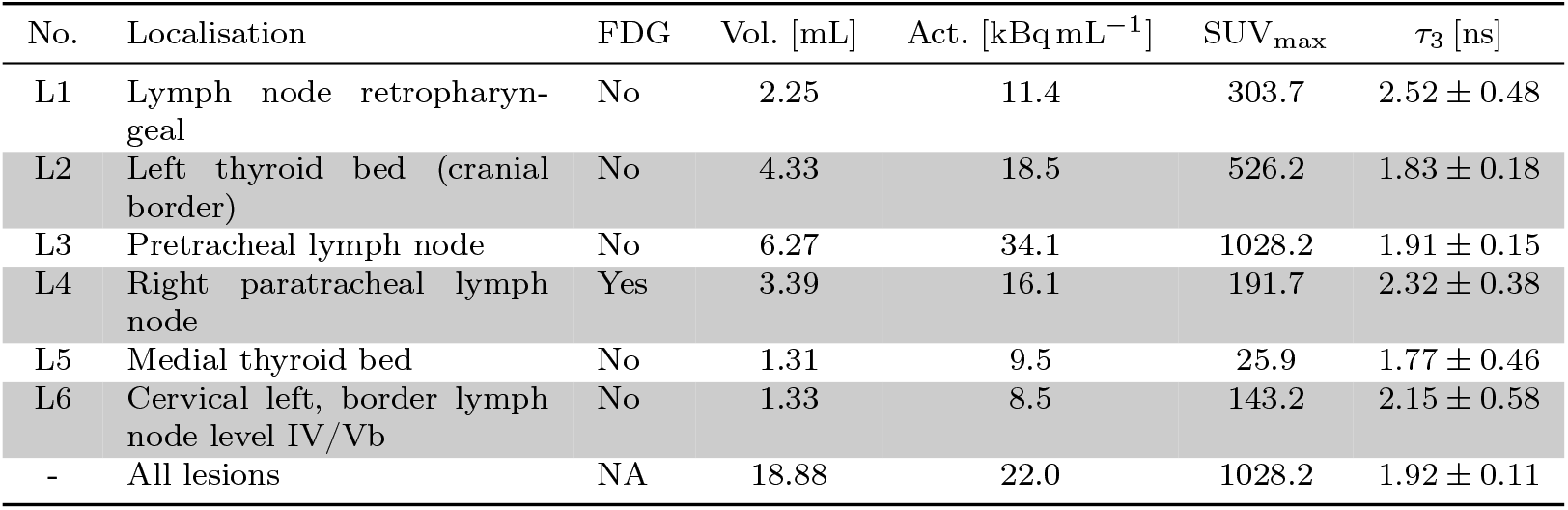
Description and oPs lifetimes in S1’s lesions. The activity from the ^124^I coincidence scan is the mean voxel value in the lesions.

| No. | Localisation | FDG | Vol. [mL] | Act. [$\text{kBq mL}^{-1}$] | $\text{SUV}_{\text{max}}$ | $\tau_3$ [ns] |
| --- | --- | --- | --- | --- | --- | --- |
| L1 | Lymph node retropharyngeal | No | 2.25 | 11.4 | 303.7 | $2.52 \pm 0.48$ |
| L2 | Left thyroid bed (cranial border) | No | 4.33 | 18.5 | 526.2 | $1.83 \pm 0.18$ |
| L3 | Pretracheal lymph node | No | 6.27 | 34.1 | 1028.2 | $1.91 \pm 0.15$ |
| L4 | Right paratracheal lymph node | Yes | 3.39 | 16.1 | 191.7 | $2.32 \pm 0.38$ |
| L5 | Medial thyroid bed | No | 1.31 | 9.5 | 25.9 | $1.77 \pm 0.46$ |
| L6 | Cervical left, border lymph node level IV/Vb | No | 1.33 | 8.5 | 143.2 | $2.15 \pm 0.58$ |
| - | All lesions | NA | 18.88 | 22.0 | 1028.2 | $1.92 \pm 0.11$ |

**Table 2:** Fit results for all early scan VOI.

| VOI | $\tau_3$ [ns] | $BR_1$ | $HDI_{BR_1}$ | $BR_2$ | $HDI_{BR_2}$ | $BR_3$ | $HDI_{BR_3}$ |
| --- | --- | --- | --- | --- | --- | --- | --- |
| Left atrium S1 | $1.78 \pm 0.28$ | 0.01 | [0.0, 0.01] | 0.72 | [0.7, 0.75] | 0.27 | [0.25, 0.29] |
| Left atrium S2 | $1.3 \pm 0.19$ | 0.01 | [0.0, 0.01] | 0.71 | [0.69, 0.74] | 0.28 | [0.26, 0.3] |
| Left atrium S3 | $1.83 \pm 0.17$ | 0.01 | [0.0, 0.01] | 0.77 | [0.76, 0.78] | 0.22 | [0.21, 0.23] |
| Right atrium S1 | $1.57 \pm 0.28$ | 0.01 | [0.0, 0.01] | 0.66 | [0.64, 0.7] | 0.33 | [0.3, 0.35] |
| Right atrium S2 | $1.76 \pm 0.16$ | 0.01 | [0.0, 0.01] | 0.75 | [0.74, 0.77] | 0.24 | [0.23, 0.25] |
| Right atrium S3 | $1.79 \pm 0.13$ | 0.01 | [0.0, 0.01] | 0.76 | [0.75, 0.77] | 0.23 | [0.22, 0.24] |
| Left parotid gland S1 | $1.92 \pm 0.13$ | 0.13 | [0.12, 0.14] | 0.6 | [0.58, 0.62] | 0.27 | [0.26, 0.28] |
| Left parotid gland S2 | $2.64 \pm 0.4$ | 0.1 | [0.08, 0.13] | 0.62 | [0.59, 0.66] | 0.27 | [0.26, 0.29] |
| Left parotid gland S2 | $2.64 \pm 0.4$ | 0.1 | [0.08, 0.13] | 0.62 | [0.59, 0.66] | 0.27 | [0.26, 0.29] |
| Right parotid gland S1 | $2.24 \pm 0.12$ | 0.08 | [0.07, 0.09] | 0.65 | [0.64, 0.66] | 0.27 | [0.27, 0.28] |
| Right parotid gland S2 | $2.25 \pm 0.26$ | 0.03 | [0.02, 0.05] | 0.71 | [0.69, 0.74] | 0.25 | [0.24, 0.27] |
| Right parotid gland S2 | $2.25 \pm 0.26$ | 0.03 | [0.02, 0.05] | 0.71 | [0.69, 0.74] | 0.25 | [0.24, 0.27] |
| Left kidney S1 | $1.37 \pm 0.28$ | 0.01 | [0.0, 0.01] | 0.62 | [0.59, 0.68] | 0.37 | [0.32, 0.4] |
| Left kidney S2 | $1.92 \pm 0.18$ | 0.0 | [0.0, 0.0] | 0.72 | [0.71, 0.74] | 0.27 | [0.26, 0.28] |
| Left kidney S3 | $1.87 \pm 0.12$ | 0.01 | [0.0, 0.01] | 0.75 | [0.74, 0.76] | 0.25 | [0.24, 0.26] |
| Right kidney S1 | $1.32 \pm 0.18$ | 0.01 | [0.0, 0.01] | 0.61 | [0.58, 0.64] | 0.38 | [0.35, 0.41] |
| Right kidney S2 | $1.6 \pm 0.14$ | 0.01 | [0.0, 0.01] | 0.71 | [0.7, 0.73] | 0.28 | [0.27, 0.3] |
| Right kidney S3 | $1.96 \pm 0.13$ | 0.01 | [0.0, 0.02] | 0.74 | [0.73, 0.76] | 0.25 | [0.24, 0.25] |
| Liver S1 | $1.53 \pm 0.12$ | 0.01 | [0.0, 0.01] | 0.7 | [0.68, 0.71] | 0.3 | [0.28, 0.31] |
| Liver S2 | $1.68 \pm 0.08$ | 0.0 | [0.0, 0.0] | 0.74 | [0.73, 0.74] | 0.26 | [0.26, 0.27] |
| Liver S3 | $1.78 \pm 0.07$ | 0.01 | [0.0, 0.0] | 0.74 | [0.74, 0.75] | 0.25 | [0.25, 0.26] |
| Spleen S1 | $1.28 \pm 0.16$ | 0.01 | [0.0, 0.01] | 0.64 | [0.62, 0.68] | 0.35 | [0.32, 0.37] |
| Spleen S2 | $1.61 \pm 0.12$ | 0.01 | [0.0, 0.01] | 0.73 | [0.72, 0.75] | 0.26 | [0.25, 0.27] |
| Spleen S3 | $1.68 \pm 0.09$ | 0.01 | [0.0, 0.01] | 0.75 | [0.74, 0.76] | 0.24 | [0.24, 0.25] |

Finally, the bottom row Fig. 4 visualizes the dependence of Δ*τ*_3_ on the maximal activity concentration in voxels of 7 ×7 ×7 mm^3^ and on the mean activity contained in a VOI. For the voxel-based fit of the oPs lifetime, no significant correlation with the activity concentration from the coincidence PET can be established. However, for entire VOI it seems that the statistical precision does not improve significantly beyond an activity of 𝒪 (100 kBq).

## 4 Discussion

^124^I has been shown to be the best candidate for oPsLI on Quadra [21, 22]. On one side, the reasonably high prompt photon BR and its low energy make the 3*γ*E selection algorithm from Ref. [20] and Quadra’s detection capabilities less prone to random events. On the other side, [^124^I]Iodide has very high and specific uptake in thyroid cancer lesions, despite having a lower activity that is administered to the patients compared to e.g. ^68^Ga and ^82^Rb. Because the differentiation of thyroid cancer lesions may be linked to oxygenation levels (see e.g. Ref. [31]), it could be relevant to utilize oPs lifetime measurements for a characterization of thyroid cancer lesions.

As in previous studies on in vivo oPs lifetime measurements, count statistics is the main challenge [19, 21]. In order to distinguish oxygenation levels in tissue through the oPs lifetime, as e.g. investigated in Ref. [5], a high number of 3*γ*E with large time differences ≳ 2 ns needs to be detected with a low random event rate. This is difficult to achieve with the current methodology under clinical conditions with respect to radionuclide, tracer, biokinetics, scan time, etc. Under conditions that are as good as it can possibly get from a clinical perspective (high uptake in S1’s lesions), our study shows that voxel-wise determination of oPs lifetimes remains challenging. The right columns in Fig. 1 shows that even for the relatively large voxel size of 7 ×7× 7 mm^3^ the error on *τ*_3_ is too large to draw any clinically valuable information from the oPsLI. Longer scan time and/or higher administrated activities would improve statistics, which are however not fesable in routine clinical practice. For the PALS in Fig. 1, only the voxel with with the highest number of 3*γ*E shows reasonable statistics and indeed the statistical error on *τ*_3_ is only 14%. However, already for the “second best” voxel, the statistical error on *τ*_3_ increases by almost a factor of two. In comparison, the activity concentration in the phantom measurements from Ref. [22] was about double the activity concentration we found in S1 at the maximal spot (half of the scan time). The limiting factor with respect to oPsLI in this study is the size of the regions with activity concentrations of 𝒪 (200 kBq mL^−1^).

There does not seem to be a clear correlation between the max. activity concentration in a voxel with the error on the oPs lifetime (see bottom left panel in Fig. 4). Likely this is because the activity concentration may correlate with the total number of 3*γ*E, but not with the number of 3*γ*E with large time differences, i.e. those events that dominate the fit of *τ*_3_. Also, at the voxel level we are much more prone to mislocalizations of 3*γ*E. There may be also a significant uncertainty on the quantification in the coincidence PET image. As shown in Ref. [35], quantification of ^124^I in small volumes is strongly affected by the suboptimal positron range correction of standard reconstruction.

If we measure the oPs lifetime at the level of single VOI rather than voxels, the statistical error on *τ*_3_ drops significantly, compared to most voxels in the oPsLI. Though we do not reach the precision of Ref. [22], L3 shows a fairly good count statistics, as can be seen also in the PALS of Fig. 3. As can be seen from Fig. 3, the count statistics are reasonably good for L2 and L3. This is result is remarkable, given the small volume of these lesions. It should be noted that the prior on Δ*τ*_3_ is 0.8 ns, which means that even for L1 and L5 there is some gain of information through the data. Of course, considering all 3*γ*E in the five lesions improves the count statistics even further, as can be seen in the top panel of Fig. 4.

The results from Tab. 2 for the VOI-based fitting show that the oPs lifetimes are consistent with the lifetime in water within the statistical uncertainty (see e.g. Ref. [36]). *τ*_3_ for the parotid glands seem to be consistently higher than for other organs, though still not really statistically significant. The VOI-based error on *τ*_3_ shows a slight negative correlation with increasing mean activity in a VOI (see bottom right panel of Fig. 4). The correlation coefficient is *r* = ™0.48 (95% credible interval is [™0.72, *™*0.15]) with a Bayes factor of *BF*_10_ = 7.88. The spread of the points is still rather large, though. As discussed before, the mean activity might still not be the ideal surrogate quantity for the number of 3*γ*E with large time differences above the background of random events.

## 5 Conclusions

The physical and biokinetic properties of ^124^I make it the prime candidate for investigating oPsLI under clinical conditions with LAFOV-PET/CT and the current methodology. For the first time, we showed the feasibility of oPs lifetime measurements in humans with ^124^I. oPsLI of thyroid cancer patients can be achieved for the total body, but the 3*γ*E count statistics remains a major challenge. Determining the oPs lifetime at the VOI level is practicable, though for some lesions the activity concentration and/or volume are on the lower end. We hope that our study paves the way for more extensive studies for characterizing the differentiation of thyroid cancer metastases through oPs lifetime measurements, in particular using improved methodologies for 3*γ*E selection and image reconstruction.

## Acknowledgements

We would like to thank Ursula Auderset, Janneke Henniphof, Angela Mendes, Rosanna Rubin, Franziska Strunz and Marco Viscione for helping with the treatment planing and patient handling.

## Declarations

### Ethics approval

This study was performed in line with the principles of the Declaration of Helsinki. All patients provided written informed consent for the use of anonymized imaging data for research purposes.

### Data availability

The datasets generated and/or analyzed during the current study are available from the corresponding author on reasonable request.

### Consent to participate

Informed consent was obtained from all individual participants included in the study.

### Authors’ contributions

LM is the primary writer of the manuscript, contributed to the data acquisition and performed the data analysis. WS wrote the list-mode processing software, analyzed the data and contributed to the licensing of software for the data acquisition. TL contributed to the regulatory and practical procedures for the administration of ^124^I in Switzerland. MA and CB are responsible for the segmentation. MC and HS contributed to the licensing of software for the data acquisition. AFC performed the data acquisition. AFC, MA, SW and RS treated the patients. LM, AR und RS conceptualized the study and contributed to the interpretation of the results. All other authors helped in some capacity. All authors read and approved the final manuscript.

